# Heat and Health Service Use: A Spatiotemporal Analysis

**DOI:** 10.1101/2025.07.12.25330649

**Authors:** Rafal Chomik, Shona Bates, Cybele Dey, Chris Dietz, Jialing Lin, Limin Mao, Ben Newell, Braulio Mark Valencia Arroyo, Patricia Davidson

## Abstract

**Background:** While acute heatwaves are known to affect hospital presentations, less is known about broader health service use, regional disparities, or the effects of chronic climatic heat.

**Methods:** We analysed 20 years of weekly health service use data between 2002 and 2022 across all Australian Statistical Areas Level 4 (SA4s) geographies for four major health services: medical services (MBS), pharmaceuticals (PBS), hospital admissions, and emergency department (ED) presentations – by clinical category (cardiovascular, respiratory, mental health, other). Three population-weighted weather metrics were measured: weekly maximum temperature, heatwave events, and long-term climatic averages. Fixed-effects panel models were used to estimate short-term impacts of weather metrics on health service use while cross-sectional models were used to assess chronic exposure effects.

**Findings:** Acute exposure to dry temperature heatwaves was consistently associated with increased ED and inpatient presentations for mental health, cardiovascular, and other conditions. Apparent temperature heatwaves, incorporating humidity, showed more variable effects. MBS consultations rose modestly, while PBS dispensing declined during heat events. Long-term exposure to higher average temperatures was linked to increased inpatient care even after adjusting for socioeconomic status, remoteness, and health workforce. Disadvantaged regions experienced amplified effects, while areas with higher outdoor work exposure showed suppressed mental health care use.

**Interpretation:** Both acute and chronic heat exposures influenced health service demand across services and geographies during the study period. Heatwaves exacerbate existing healthcare inequalities and can disrupt access to medicines. Adaptation strategies must extend beyond hospitals to include primary and community-based care, with attention to population vulnerability and regional climate.

**Funding:** The International Centre for Future Health Systems receives funding from The Ian Potter Foundation.

## 1. Introduction

Climate heating poses a well-recognised public health threat (IPCC 2022; Romanello et al. 2023). While the links between heatwaves and mortality and morbidity are well established (D’Ippoliti et al. 2010; Varghese et al. 2020; Vicedo-Cabrera et al. 2021), there is growing interest in how both ambient and extreme heat influence patterns of health service use (Mason et al. 2022; Xu et al. 2023a). Understanding these effects is needed for adequate climate-adaptive health system planning.

International and Australian studies consistently report increased emergency and hospital demand during extreme heat, particularly for cardiovascular, respiratory, and mental health conditions (Xu et al. 2023b; Tong et al. 2024). However, a systematic review by Mason et al. (2022) found that most Australian research has focused on single cities, short-term exposures, and hospital-based services. Few studies have examined impacts on primary care or medication dispensing. One national analysis of GP visits during heatwaves has been reported in abstract form (Varghese et al. 2021), but no published study to date has examined nationwide impacts across all major health service types, including Medicare-subsidised consultations, pharmaceuticals, and hospital care.

This study addresses these gaps using two decades of weekly data across all Australian Statistical Areas Level 4 (SA4s) to examine how temperature affects use of four major health services: Medicare-subsidised general practice and specialist consultations (MBS), prescription pharmaceuticals (PBS), public hospital admissions, and emergency department (ED) presentations. Specifically, we ask: (1) How does weekly heat exposure affect utilisation of health services across clinical conditions? (2) What is the impact of extreme heat thresholds, such as days above 35°C and heatwave events? and (3) Do areas with hotter long-term climates exhibit higher average service use, independent of socioeconomic, remoteness, and demographic factors? We also examine whether these associations vary by geography, disadvantage, and occupational exposure to outdoor work.

## 2. Data and methods Study design

We conducted a spatiotemporal ecological analysis using 20 years of weekly data from all Australian Statistical Areas Level 4 (SA4s), which represent large labour market and health service regions defined by the Australian Bureau of Statistics. The study period spanned 2002 to 2022, and the unit of analysis was the SA4 week. (ABS, 2021; see Figure 1).

**Figure 1.**
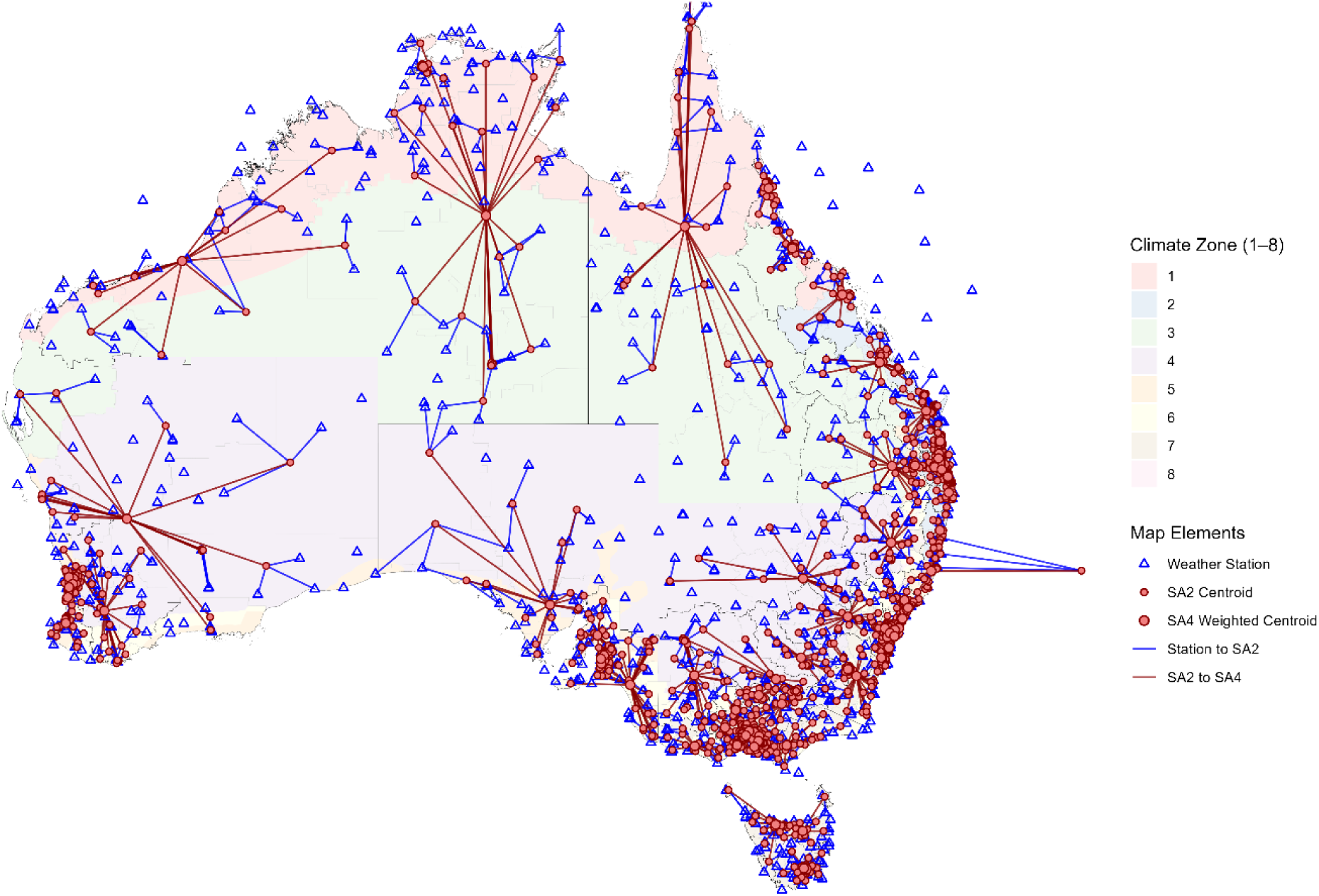
Weather station to SA2 to SA4 flow map with climate zones. Notes: Climate zones based on Australian Building Codes Board categories.

### Health service use data

Health services data was sourced from AIHW (2024), with SA4-level hospitalisations (2002 to 2022), ED presentations (2014 to 2022), MBS claims (2002 to 2022), and PBS dispensations (2002 to 2022). Condition-specific service use records from each dataset were mapped to four clinical groupings – Cardiovascular, Respiratory, Mental health, and Other – in line with established specifications (AIHW 2024).

### Weather data

Population weighted weather variables were derived from station data and aggregated to the SA4-week level (Figure 1). These included weekly: (i) average maximum temperature; (ii) average apparent temperature, calculated using the Steadman formula to account for humidity (Steadman, 1979); (iii) number of days over 35°C; (iv) number of apparent temperature heatwave days; and (v) number of dry temperature heatwave days.

In this preliminary analysis, heatwave days were defined using an adapted version of the EHF framework (Nairn & Fawcett 2015), in which a heatwave is identified when the 3-day average maximum temperature exceeds both a 30-day rolling acclimatisation baseline and a 15-day seasonal climatological norm. Heatwave days correspond to periods where the EHF is greater than zero. In using *daily maximum temperature*, rather than daily mean temperature, to calculate EHF for both dry and apparent temperature, the current iteration ignores night-time temperature. A future version of this analysis will incorporate daily mean temperature and will test severity classifications using the 85th percentile of EHF values, as well as absolute temperature thresholds (e.g. 3-day means above 30 °C or maximum temperatures above 35 °C) to better reflect physiologically meaningful heat exposure.

### Area characteristics data

Six area-level covariates were included in models of long-term exposure: (i) socioeconomic status, measured using the 2021 relative percentile rank of the Index of Relative Socioeconomic Advantage and Disadvantage (IRSAD), from the Australian Bureau of Statistics; (ii) remoteness, based on the 2019 Modified Monash Model (MMM7), which incorporates both remoteness and town size to capture service accessibility; (iii) the proportion of the population aged 65 years and over based on the 2021 Census; (iv) chronic illness prevalence based on the 2021 Census; (v) health workforce availability, drawn from the 2021 National Health Workforce Dataset and matched to service type (i.e. medical practitioners for MBS, pharmacists for PBS, nurses and midwives for hospitals and EDs); and (vi) outdoor work exposure, derived from mapping US O*NET occupational heat stress scores onto to the Australian context using concordance tables and weighted by 2021 Census employment data (DESE 2020; Tuccio et al. 2023; Chomik 2025).

### Statistical analysis

Short-term impacts were estimated using fixed-effects linear panel regression models with a SA4–week structure. The outcome was the crude weekly rate of service use per 100,000 population. Exposure variables included average weekly maximum and apparent temperature, number of days above 35 °C, and number of heatwave days (based on dry and apparent temperature). Models included fixed effects for SA4, week-of-year, and year. Interaction terms tested whether effects varied by socioeconomic status or outdoor work exposure. A future iteration will revise exposures to use daily mean temperature rather than maximum temperature alone, and incorporate both heatwave severity and absolute thresholds more relevant to physiological heat stress.

Long-term associations were assessed using cross-sectional linear models, with the mean weekly crude rate of service use as the outcome and long-term average maximum temperature as the main exposure. Models were adjusted for all area-level covariates. Future versions will replace the exposure variable with long-term average daily mean temperature, to better reflect cumulative and night-time heat. Analyses were conducted in R.

## 3. Results Descriptive statistics

Over the two decades, all Australian SA4s experienced both dry and apparent temperature heatwaves, averaging 120–150 dry and 130–160 apparent heatwave days per year. Days over 35°C ranged from fewer than one annually in temperate regions to over 160 in parts of inland Queensland.

Health service use showed clear seasonal variation (Figure 2). Respiratory hospitalisations peaked in late August, while mental health-related MBS use was highest between December and March. MBS and PBS services for cardiovascular and respiratory conditions were relatively stable year-round. Hospital admissions declined around Christmas.

**Figure 2.**
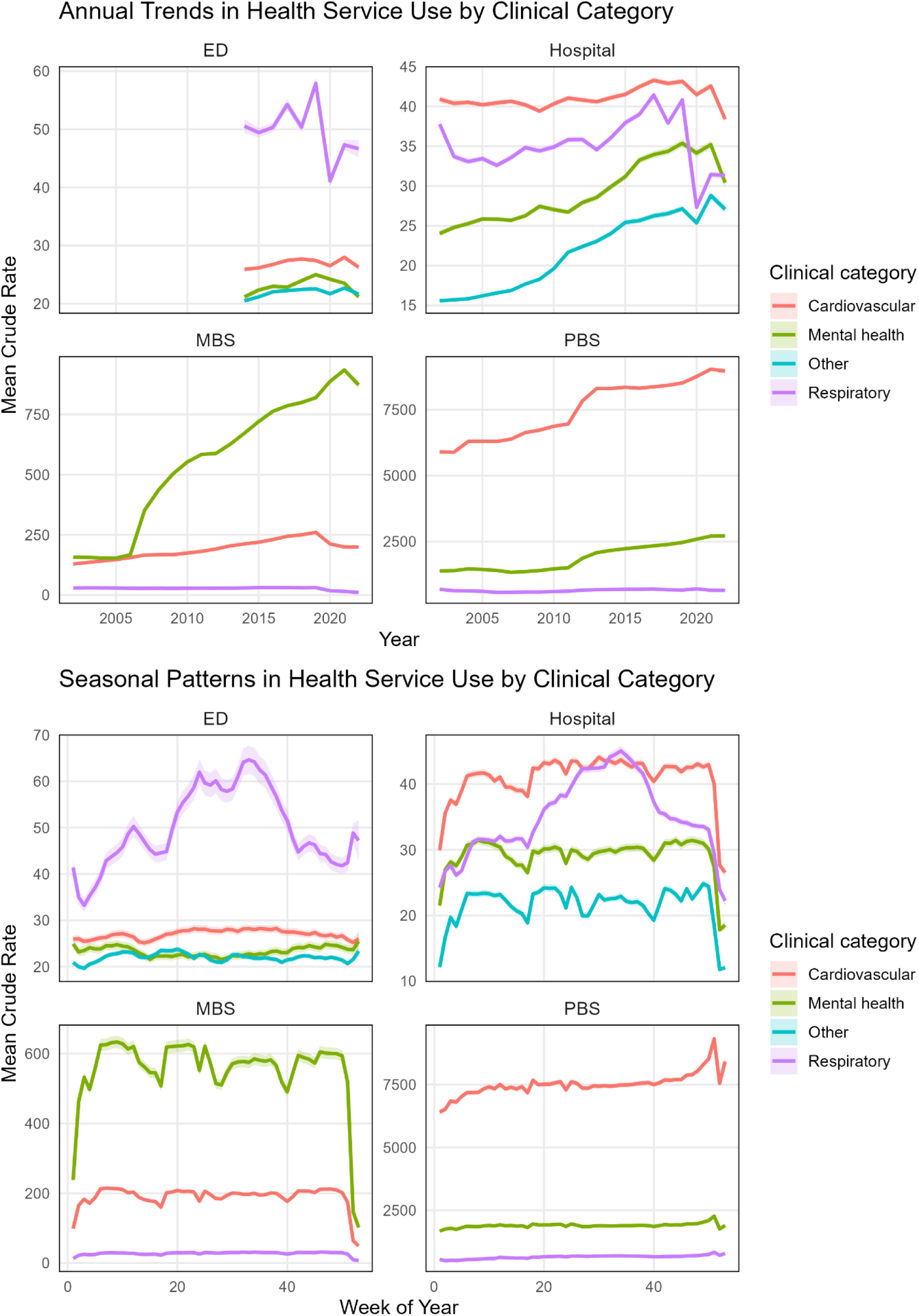
Health service use annual and seasonal trends by clinical category. Source: Authors’ analysis of AIHW data

National trends over time varied by condition and service. The most marked change was a steep rise in MBS mental health consultations following the Better Access initiative in 2006 (Meadows et al. 2015), increasing from 157 to 873 per 100,000 between 2002 and 2022. PBS dispensing for mental health also nearly doubled, especially after 2011. In contrast, respiratory-related MBS, PBS, and hospital services declined modestly, with a sharper drop during the COVID-19 pandemic, likely due to reduced influenza transmission (Sullivan et al. 2020; Australian Government 2021).

ED presentations remained stable across conditions, while hospital admissions rose for mental health (+26%) and other conditions (+75%), consistent with broader health system reforms and evolving clinical practice (Pirkis et al. 2011; Department of Health 2012).

### Short term acute heat exposure

The most consistent evidence of increased health service use came from dry temperature heatwaves, as currently defined (Figure 3). These events were associated with higher rates of ED and hospital care. Each additional dry temperature heatwave day was linked to +0.11 ED presentations and +0.21 hospitalisations for mental health per 100,000 population (both p < 0.001). Cardiovascular and total hospitalisations also rose significantly (+0.03 and +0.22, both p < 0.01).

**Figure 3.**
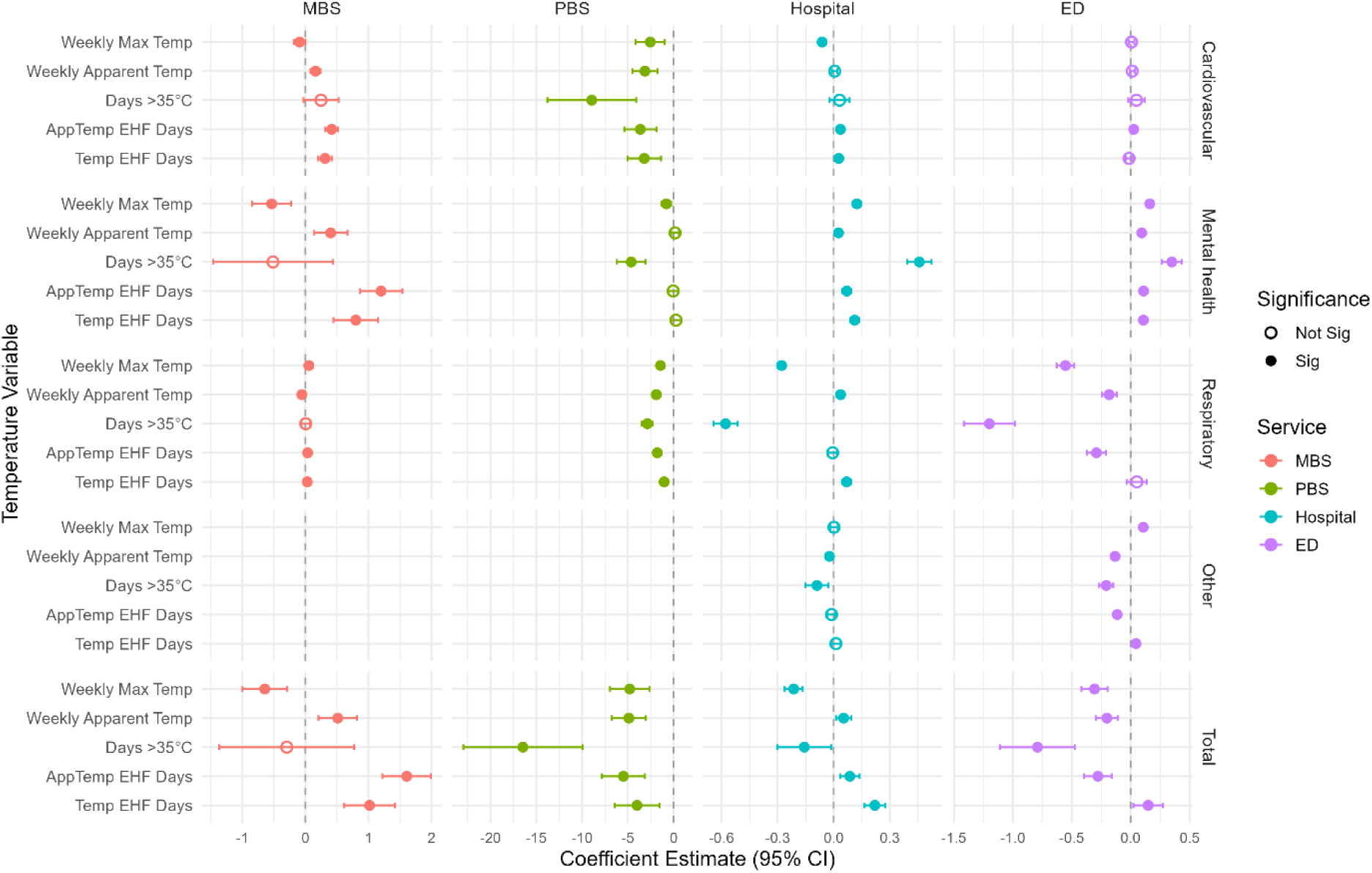
Short term heat exposure and health service use. Figure note: Coefficients are from linear models at the SA4 level, standardised for comparability. Full model results, including all predictors and unstandardised estimates, are available in Supplementary Tables

MBS care also increased, particularly for mental health and cardiovascular conditions. Each dry temperature heatwave day was associated with +0.80 MBS mental health consultations and +0.31 cardiovascular consultations (both p < 0.001). In contrast, pharmaceutical dispensing declined, including –3.21 PBS items for cardiovascular, – 1.04 for respiratory, and –4.00 for total conditions (all p < 0.01).

Other heat indicators showed more variable effects. Weekly maximum temperature was associated with increased MBS care (+0.40, p < 0.01) and ED use (+0.16, p < 0.001) per 1°C increase, but hospital-based respiratory care declined (–0.55 ED and – 0.28 hospital, both p < 0.001). Apparent temperature heatwaves were also linked to greater mental health service use, including +1.20 MBS consultations, +0.07 hospitalisations, and +0.11 ED presentations per day (all p < 0.001), and a smaller rise in cardiovascular MBS use (+0.42, p < 0.001). However, these events were associated with declines in total ED use (–0.28 per day, p < 0.001) and PBS dispensing across conditions.

Stratified models by jurisdiction and climate zone revealed consistent patterns. Dry temperature heatwaves were more strongly associated with increased ED and hospital use across most states and territories, particularly for mental health. MBS use also rose in many areas, especially for mental health and cardiovascular care. By climate zone, Hot Dry and Temperate/Cold regions showed the strongest associations between dry temperature heatwaves and health service use. In contrast, apparent temperature heatwaves showed clearer effects in Hot Humid zones, particularly for mental health-related MBS and ED visits, though associations were generally smaller and less consistent elsewhere.

Interaction models showed that disadvantaged areas experienced amplified effects of heatwaves on mental health and hospital care, alongside reduced pharmaceutical dispensing. In contrast, areas with higher outdoor work exposure showed reduced mental health service use during heatwaves, including lower MBS consultations and hospitalisations, while PBS dispensing modestly increased.

### Chronic Heat Exposure

Long-term exposure to higher average weekly maximum temperatures was associated with increased ED and hospital presentations, even after controlling for covariates such as socioeconomic status, remoteness, and health access (Figure 4). In the main models a 1°C increase in long-term average weekly (dry bulb) maximum temperature was linked to +0.40 cardiovascular, +0.29 respiratory, +0.25 mental health, and +0.41 other-condition ED presentations per 100,000 people per week (all p < 0.01). Hospital admissions also increased with temperature, though with smaller magnitudes (e.g. +0.22 cardiovascular and +0.27 respiratory, both p < 0.01).

**Figure 4.**
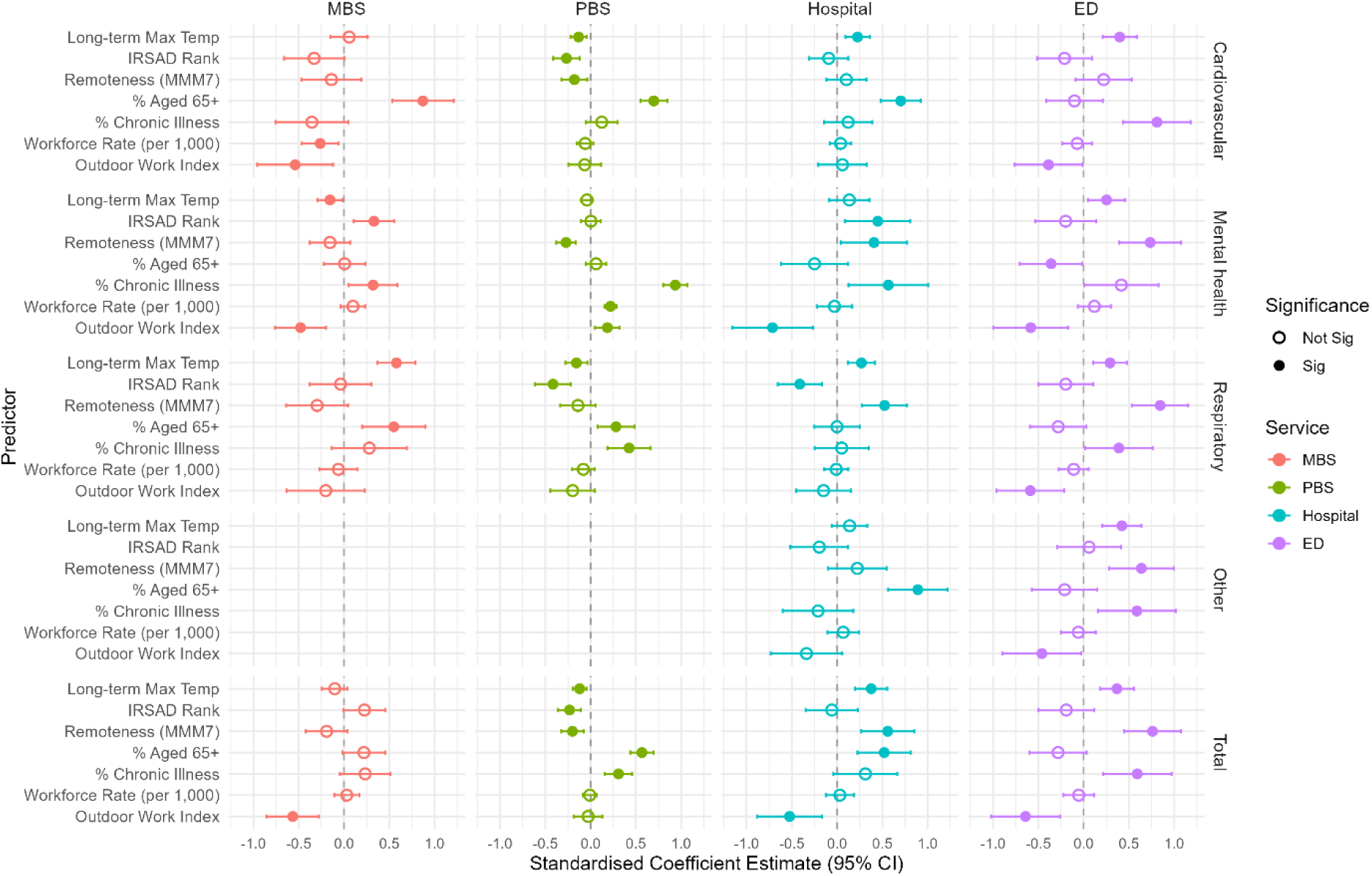
Long term heat exposure and health service use. Figure note: Coefficients are from linear models at the SA4 level, standardised for comparability. Full model results, including all predictors and unstandardised estimates, are available in Supplementary Tables

Associations with MBS and PBS use were more variable. Respiratory-related MBS consultations rose with higher temperatures (+0.56, p < 0.001), but cardiovascular and mental health MBS use showed weaker or non-significant associations. Pharmaceutical dispensing (PBS) was negatively associated with long-term temperature, particularly for cardiovascular (–0.12, p < 0.01) and respiratory conditions (–0.15, p < 0.05).

Covariates generally behaved as expected. Higher proportions of older adults and those with chronic illness were associated with greater hospital service use. Socioeconomic disadvantage and remoteness were associated with lower levels of care typically provided by GPs across most conditions, while ED use for respiratory, mental health was higher in disadvantaged or remote areas. Health workforce availability showed mixed or non-significant effects.

The outdoor work index was negatively associated with service use in fully adjusted models, particularly ED visits (–4.54, p = 0.003). However, the effect weakened or reversed when socioeconomic or remoteness covariates were removed. When both were excluded, the association became positive (+1.70, p = 0.089).

Analyses restricted to the post-2018 period, including the COVID-19 pandemic, yielded consistent results. In some cases, associations strengthened, particularly between long-term maximum temperature and hospitalisations for cardiovascular and respiratory illness.

## 4. Discussion

This study finds that short-term temperature extremes, particularly dry temperature heatwaves, are associated with increased health service use across Australia. Mental health care appeared especially heat-sensitive, consistent with previous studies linking high temperatures to increased mental health presentations (Xu et al. 2023a; Nori-Sarma et al. 2022). In contrast, dispensing of pharmaceutical items declined during heat events, potentially reflecting reduced access or reprioritisation of care. Apparent temperature heatwaves, as currently defined, also affected service use, though less consistently, suggesting that humidity-related stress may be more relevant in some climates than others. This finding may be sensitive to relative measure of heatwaves which will be tested in future iterations of this analysis. The findings extend the Australian evidence base beyond city- and hospital-based studies (Mason et al. 2022), showing that diverse services are affected across geographic settings.

Higher long-term average temperatures were associated with greater use of hospital and ED services, but lower use of Medicare-subsidised care and pharmaceuticals. These results align with studies highlighting structural differences in service availability, housing conditions, and climate adaptation across regions (Thomson et al. 2023; Amoatey et al. 2025). As these models reflect multi-year averages, the associations likely capture the evolved structural geographic disparities in exposure and healthcare access.

Heat-related service impacts were not limited to the most heat-prone or remote regions; temperate and subtropical areas also showed strong associations. In Hot Humid zones, apparent temperature effects were more pronounced, aligning with known physiological responses to combined heat and humidity (Borg et al. 2019). Socioeconomically disadvantaged areas experienced greater increases in mental health and hospital service use and sharper declines in dispensing, consistent with prior work identifying vulnerability to heat-related health risks (Tong et al. 2024; Xu et al. 2023b). In regions with higher outdoor work exposure, health service use during heat was lower, possibly reflecting differences in health-seeking behaviour shaped by occupational culture, gender norms, or access barriers. When socioeconomic and remoteness controls were removed, the association reversed, suggesting that outdoor work may proxy for patterns of suppressed demand rather than lower heat-related need.

These findings reinforce calls for climate-adaptive health systems that address both acute and chronic exposures. The Australian Government’s National Health and Climate Strategy (2023) highlights the need for localised, evidence-informed responses. Our results suggest this must include not only emergency care, but also general practice, pharmaceutical services, and community-level planning, particularly in vulnerable areas. Even a 0.5 °C increase in global temperatures could substantially affect system demand (King et al. 2017), underscoring the importance of anticipatory action (Tait et al. 2018).

The analysis has limitations. Current exposure variables may not fully capture physiologically meaningful events. EHF was calculated using daily maximum rather than mean temperature; future work will adopt standard methods and include heatwave severity and absolute temperature thresholds. We will also test lagged exposure models to assess whether declines in pharmaceutical dispensing reflect temporary delays or more persistent disruption.

Our short term exppsure models estimate the average marginal effect per heatwave day and do not distinguish between isolated and consecutive heat days. While some studies suggest that single hot days can be as impactful as multi-day heatwaves (Dey et al. 2025), particularly when defined by absolute thresholds, EHF-based analyses typically focus on multi-day events. Future work could explore whether effects accumulate nonlinearly or differ between isolated and clustered heat days.

Regional variation in heat sensitivity likely reflects differences in access to cooling, housing conditions, and behavioural adaptation. While our fixed-effects models estimate average within-region associations and test some interactions, future work will explore using further interaction terms or random slope models to test spatial variation in heat response and identify high-risk communities.

While our chronic exposure models used long-term average maximum temperature, this may understate the impact of heat extremes. Future iterations could incorporate cumulative exposure metrics, such as annual counts of days above 35 °C or EHF days, better reflecting heat intensity and frequency. For example, recent work by He et al. (2024) linked the mental health burden to annual counts of hot days, reinforcing the importance of event-based metrics for long-term adaptation planning.

Remaining limitations are based on data availability, including that crude rates may mask age-related variation while weekly groupings and large geographic areas in the case of regional SA4s may mask more granular differences and may not align fully to weather data. A future iteration may test robustness by using alternative weather data (i.e., Bureau’s Atmospheric high-resolution Regional Reanalysis for Australia).

## 5. Conclusion

This study demonstrates that both acute and chronic heat exposures influence health service use across a broad range of settings, conditions, and geographies in Australia. Heatwaves were consistently associated with increased ED and hospital presentations, especially for mental health, while apparent temperature heatwaves showed more variable effects. Care typically delivered by general practitioners also rose during hot weeks, but pharmaceutical dispensing declined, suggesting that access may be disrupted during extreme heat events.

## Data Availability

All data produced in the present study are available upon reasonable request to the authors

## Notes

### Competing Interest Statement

The authors have declared no competing interest.

