## Supplemental tables for "Heat and Health Service Use: A Spatiotemporal Analysis"

### Supplementary Table 1: Full fixed-effects model results (national)

| condition | term | MBS | PBS | Hospital | ED |
| --- | --- | --- | --- | --- | --- |
| Cardiovascular | avg_weekly_max | -0.092 () | -2.557 (**) | -0.062 (***) | 0.008 () |
| Cardiovascular | avg_weekly_apptemp | 0.159 (***) | -3.136 (***) | 0.005 () | 0.014 () |
| Cardiovascular | numb_days_over35 | 0.247 () | -8.946 (***) | 0.031 () | 0.049 () |
| Cardiovascular | numb_apptemp_hw_days | 0.416 (***) | -3.642 (***) | 0.036 (***) | 0.026 (*) |
| Cardiovascular | numb_temp_hw_days | 0.311 (***) | -3.21 (**) | 0.027 (**) | -0.014 () |
| Mental health | avg_weekly_max | -0.536 (**) | -0.801 (**) | 0.124 (***) | 0.162 (***) |
| Mental health | avg_weekly_apptemp | 0.4 (**) | 0.147 () | 0.026 (**) | 0.094 (***) |
| Mental health | numb_days_over35 | -0.516 () | -4.643 (***) | 0.458 (***) | 0.35 (***) |
| Mental health | numb_apptemp_hw_days | 1.201 (***) | -0.056 () | 0.07 (***) | 0.109 (***) |
| Mental health | numb_temp_hw_days | 0.799 (***) | 0.248 () | 0.112 (***) | 0.108 (***) |
| Respiratory | avg_weekly_max | 0.054 (***) | -1.439 (***) | -0.278 (***) | -0.553 (***) |
| Respiratory | avg_weekly_apptemp | -0.056 (***) | -1.898 (***) | 0.037 (***) | -0.182 (***) |
| Respiratory | numb_days_over35 | 0.004 () | -2.875 (***) | -0.578 (***) | -1.199 (***) |
| Respiratory | numb_apptemp_hw_days | 0.034 (**) | -1.786 (***) | -0.006 () | -0.29 (***) |
| Respiratory | numb_temp_hw_days | 0.028 (*) | -1.041 (***) | 0.069 (***) | 0.053 () |
| Total | avg_weekly_max | -0.646 (***) | -4.797 (***) | -0.214 (***) | -0.307 (***) |
| Total | avg_weekly_apptemp | 0.512 (**) | -4.887 (***) | 0.053 (*) | -0.202 (***) |
| Total | numb_days_over35 | -0.296 () | -16.464 (***) | -0.157 (*) | -0.79 (***) |
| Total | numb_apptemp_hw_days | 1.607 (***) | -5.485 (***) | 0.086 (**) | -0.278 (***) |
| Total | numb_temp_hw_days | 1.018 (***) | -4.003 (**) | 0.22 (***) | 0.149 (*) |
| Other | avg_weekly_max |  |  | 0.001 () | 0.107 (***) |
| Other | avg_weekly_apptemp |  |  | -0.023 (*) | -0.132 (***) |
| Other | numb_days_over35 |  |  | -0.09 (**) | -0.207 (***) |
| Other | numb_apptemp_hw_days |  |  | -0.012 () | -0.114 (***) |
| Other | numb_temp_hw_days |  |  | 0.012 () | 0.045 (***) |

### Supplementary Table 2: Jurisdiction-stratified model results

| jurisdiction | condition | temp_var | MBS | PBS | Hospital | ED |
| --- | --- | --- | --- | --- | --- | --- |
| ACT | Cardiovascular | avg_weekly_max | -0.162 () | 7.21 () | 0.044 () | 0.302 (*) |
| ACT | Cardiovascular | avg_weekly_apptemp | -0.167 () | -4.776 () | -0.016 () | -0.107 () |
| ACT | Cardiovascular | numb_days_over35 | -2.529 (**) | 27.363 () | 0.617 (**) | 0.834 (*) |
| ACT | Cardiovascular | numb_apptemp_hw_days | 0.03 () | -1.566 () | -0.022 () | -0.122 () |
| ACT | Cardiovascular | numb_temp_hw_days | 0.2 () | 1.011 () | -0.006 () | 0.156 () |
| ACT | Mental health | avg_weekly_max | -1.057 () | 3.639 (**) | -0.02 () | 0.143 (*) |
| ACT | Mental health | avg_weekly_apptemp | -0.136 () | -1.814 () | 0.056 () | 0.129 (*) |
| ACT | Mental health | numb_days_over35 | -5.424 () | 3.798 () | 0.018 () | 0.309 () |
| ACT | Mental health | numb_apptemp_hw_days | 0.368 () | -0.43 () | 0.08 () | 0.177 (**) |
| ACT | Mental health | numb_temp_hw_days | 0.362 () | 3.162 (*) | -0.015 () | 0.124 () |
| ACT | Respiratory | avg_weekly_max | 0.011 () | 0.217 () | -0.017 () | 0.105 () |
| ACT | Respiratory | avg_weekly_apptemp | -0.016 () | -0.769 () | 0.064 () | -0.28 () |
| ACT | Respiratory | numb_days_over35 | -0.115 () | 2.536 () | -0.013 () | 0.506 () |
| ACT | Respiratory | numb_apptemp_hw_days | -0.024 () | -0.971 () | 0.023 () | -0.312 () |
| ACT | Respiratory | numb_temp_hw_days | 0.016 () | -0.21 () | -0.035 () | 0.109 () |
| ACT | Total | avg_weekly_max | -1.161 () | 11.066 (*) | 0.025 () | 0.615 (*) |
| ACT | Total | avg_weekly_apptemp | -0.267 () | -7.359 () | 0.054 () | -0.401 () |
| ACT | Total | numb_days_over35 | -7.41 () | 33.697 () | 0.509 () | 1 () |
| ACT | Total | numb_apptemp_hw_days | 0.4 () | -2.967 () | 0.028 () | -0.441 () |
| ACT | Total | numb_temp_hw_days | 0.419 () | 3.964 () | -0.032 () | 0.577 (*) |
| NSW | Cardiovascular | avg_weekly_max | -0.217 (*) | 4.058 (**) | -0.077 (***) | 0.004 () |
| NSW | Cardiovascular | avg_weekly_apptemp | 0.165 (*) | -3.548 (**) | 0.028 (*) | 0.019 () |
| NSW | Cardiovascular | numb_days_over35 | -1.414 (***) | 24.672 (***) | -0.19 (**) | 0.071 () |
| NSW | Cardiovascular | numb_apptemp_hw_days | 0.324 (**) | -4.927 (**) | 0.041 (*) | 0.027 () |
| NSW | Cardiovascular | numb_temp_hw_days | 0.018 () | -2.061 () | -0.043 (*) | -0.016 () |
| NSW | Mental health | avg_weekly_max | 1.09 (***) | 0.553 () | 0.142 (***) | 0.265 (***) |
| NSW | Mental health | avg_weekly_apptemp | -0.252 () | -0.323 () | 0.033 (*) | 0.09 (**) |
| NSW | Mental health | numb_days_over35 | 2.553 (*) | 1.841 () | 0.325 (***) | 0.805 (***) |
| NSW | Mental health | numb_apptemp_hw_days | 1.388 (***) | -0.22 () | 0.109 (***) | 0.164 (***) |
| NSW | Mental health | numb_temp_hw_days | 0.962 (**) | 0.971 () | 0.084 (***) | 0.089 (*) |
| NSW | Respiratory | avg_weekly_max | -0.006 () | -1.176 (***) | -0.391 (***) | -1.087 (***) |
| NSW | Respiratory | avg_weekly_apptemp | 0.068 (**) | -1.578 (***) | 0.093 (***) | -0.088 () |
| NSW | Respiratory | numb_days_over35 | -0.135 () | -3.629 (***) | -0.733 (***) | -0.977 (***) |
| NSW | Respiratory | numb_apptemp_hw_days | 0.04 () | -2.264 (***) | 0.007 () | -0.484 (***) |
| NSW | Respiratory | numb_temp_hw_days | -0.015 () | -1.131 (***) | 0.033 () | 0.116 () |
| NSW | Total | avg_weekly_max | 1.087 (**) | 3.434 () | -0.313 (***) | -0.742 (***) |
| NSW | Total | avg_weekly_apptemp | -0.065 () | -5.449 (**) | 0.128 (***) | -0.115 () |
| NSW | Total | numb_days_over35 | 1.834 () | 22.884 (**) | -0.526 (***) | -0.359 () |
| NSW | Total | numb_apptemp_hw_days | 1.769 (***) | -7.411 (**) | 0.14 (**) | -0.463 (***) |
| NSW | Total | numb_temp_hw_days | 0.937 (*) | -2.221 () | 0.04 () | 0.261 (**) |
| NT | Cardiovascular | avg_weekly_max | 0.624 (**) | 1.132 () | -0.098 () | -0.036 () |
| NT | Cardiovascular | avg_weekly_apptemp | -0.221 () | 2.412 () | -0.133 () | 0.027 () |
| NT | Cardiovascular | numb_days_over35 | 0.889 (**) | -0.045 () | -0.142 () | 0.003 () |
| NT | Cardiovascular | numb_apptemp_hw_days | -0.052 () | -3.403 () | -0.013 () | 0.111 () |
| NT | Cardiovascular | numb_temp_hw_days | 0.346 () | 3.577 () | 0.044 () | 0.07 () |
| NT | Mental health | avg_weekly_max | 0.761 (*) | 0.123 () | 0.351 (***) | 0.65 (***) |
| NT | Mental health | avg_weekly_apptemp | 0.642 () | 0.892 () | 0.099 () | 0.17 () |
| NT | Mental health | numb_days_over35 | 0.692 () | 0.336 () | 0.392 (***) | 0.313 () |
| NT | Mental health | numb_apptemp_hw_days | 0.317 () | -0.628 () | 0.15 (*) | 0.117 () |
| NT | Mental health | numb_temp_hw_days | 1.223 (***) | 1.824 (*) | 0 () | 0.217 () |
| NT | Respiratory | avg_weekly_max | -0.035 () | -0.49 () | -1.259 (***) | -3.191 (***) |
| NT | Respiratory | avg_weekly_apptemp | 0.088 () | -0.354 () | 0.112 () | 0.707 () |
| NT | Respiratory | numb_days_over35 | -0.044 () | -0.008 () | -1.685 (***) | -3.601 (***) |
| NT | Respiratory | numb_apptemp_hw_days | 0.118 () | -0.573 (*) | 0.041 () | -0.556 () |
| NT | Respiratory | numb_temp_hw_days | -0.048 () | -0.259 () | 0.04 () | -0.254 () |
| NT | Total | avg_weekly_max | 1.482 (**) | 0.766 () | -0.886 (***) | -1.873 (**) |
| NT | Total | avg_weekly_apptemp | 0.555 () | 2.951 () | 0.132 () | 1.075 () |
| NT | Total | numb_days_over35 | 1.633 (**) | 0.282 () | -1.246 (***) | -2.719 (***) |
| NT | Total | numb_apptemp_hw_days | 0.342 () | -4.604 () | 0.217 () | -0.319 () |
| NT | Total | numb_temp_hw_days | 1.568 (***) | 5.142 () | 0.113 () | -0.085 () |
| Qld | Cardiovascular | avg_weekly_max | 0.26 (*) | -1.076 () | -0.01 () | 0.19 (***) |
| Qld | Cardiovascular | avg_weekly_apptemp | 0.477 (***) | -2.663 () | 0.027 () | 0.095 (**) |
| Qld | Cardiovascular | numb_days_over35 | 0.905 (**) | 14.652 (**) | 0.112 () | 0.202 (*) |
| Qld | Cardiovascular | numb_apptemp_hw_days | 0.601 (***) | -4.804 (**) | 0.062 (**) | 0.074 (*) |
| Qld | Cardiovascular | numb_temp_hw_days | 0.386 (***) | -2.694 () | 0.042 (*) | 0.042 () |
| Qld | Mental health | avg_weekly_max | 0.929 () | -0.376 () | 0.186 (***) | 0.149 (***) |
| Qld | Mental health | avg_weekly_apptemp | -0.128 () | -0.28 () | 0.073 (**) | 0.064 (***) |
| Qld | Mental health | numb_days_over35 | -0.525 () | -4.524 (**) | 0.287 (**) | 0.032 () |
| Qld | Mental health | numb_apptemp_hw_days | 1.112 (**) | -0.577 () | 0.114 (***) | 0.062 (**) |
| Qld | Mental health | numb_temp_hw_days | 1.453 (***) | -0.149 () | 0.116 (***) | 0.074 (**) |
| Qld | Respiratory | avg_weekly_max | -0.028 () | -3.871 (***) | -0.352 (***) | -0.214 (*) |
| Qld | Respiratory | avg_weekly_apptemp | 0.015 () | -1.861 (***) | -0.029 () | 0.017 () |
| Qld | Respiratory | numb_days_over35 | 0.191 (*) | -0.308 () | -0.896 (***) | -1.242 (***) |
| Qld | Respiratory | numb_apptemp_hw_days | 0.056 (*) | -1.952 (***) | -0.02 () | -0.232 (***) |
| Qld | Respiratory | numb_temp_hw_days | 0.098 (***) | -1.259 (***) | 0.091 (***) | 0.106 () |
| Qld | Total | avg_weekly_max | 1.245 (*) | -5.323 () | -0.181 (*) | 0.324 (**) |
| Qld | Total | avg_weekly_apptemp | 0.316 () | -4.803 (*) | 0.063 () | 0.035 () |
| Qld | Total | numb_days_over35 | 1.246 () | 9.82 () | -0.658 (***) | -0.844 (***) |
| Qld | Total | numb_apptemp_hw_days | 1.742 (***) | -7.334 (**) | 0.151 (**) | -0.26 (**) |
| Qld | Total | numb_temp_hw_days | 1.796 (***) | -4.102 () | 0.271 (***) | 0.29 (***) |
| SA | Cardiovascular | avg_weekly_max | -0.313 (*) | -10.337 (***) | -0.004 () | -0.066 () |
| SA | Cardiovascular | avg_weekly_apptemp | -0.016 () | 2.893 () | 0.07 (*) | -0.021 () |
| SA | Cardiovascular | numb_days_over35 | -1.54 (***) | -40.907 (***) | -0.073 () | -0.253 (*) |
| SA | Cardiovascular | numb_apptemp_hw_days | 0.296 () | -0.308 () | 0.04 () | -0.032 () |
| SA | Cardiovascular | numb_temp_hw_days | 0.153 () | -5.358 () | 0.004 () | -0.072 () |
| SA | Mental health | avg_weekly_max | -1.161 (**) | -2.132 (**) | 0.101 (***) | 0.118 (**) |
| SA | Mental health | avg_weekly_apptemp | -1.39 (**) | -0.501 () | 0.109 (***) | 0.087 () |
| SA | Mental health | numb_days_over35 | -4.731 (***) | -10.939 (***) | 0.203 (**) | 0.167 () |
| SA | Mental health | numb_apptemp_hw_days | 0.682 () | -0.137 () | 0.106 (***) | 0.067 () |
| SA | Mental health | numb_temp_hw_days | 0.095 () | -0.878 () | 0.112 (***) | 0.161 (**) |
| SA | Respiratory | avg_weekly_max | -0.052 () | -1.761 (***) | 0.028 () | -0.072 () |
| SA | Respiratory | avg_weekly_apptemp | 0.05 () | -1.52 (***) | -0.174 (***) | -0.04 () |
| SA | Respiratory | numb_days_over35 | -0.26 (**) | -6.935 (***) | -0.215 () | -0.336 () |
| SA | Respiratory | numb_apptemp_hw_days | 0.043 () | -1.487 (***) | -0.078 () | -0.086 () |
| SA | Respiratory | numb_temp_hw_days | 0.012 () | -0.825 () | 0.079 () | -0.109 () |
| SA | Total | avg_weekly_max | -1.391 (**) | -14.23 (***) | 0.12 () | -0.045 () |
| SA | Total | avg_weekly_apptemp | -1.238 (*) | 0.872 () | -0.054 () | -0.043 () |
| SA | Total | numb_days_over35 | -5.827 (***) | -58.78 (***) | -0.256 () | -0.825 (*) |
| SA | Total | numb_apptemp_hw_days | 0.967 () | -1.931 () | 0.012 () | -0.151 () |
| SA | Total | numb_temp_hw_days | 0.247 () | -7.062 () | 0.218 (**) | -0.022 () |
| Tas | Cardiovascular | avg_weekly_max | -0.211 () | -15.544 (**) | -0.054 () | 0.052 () |
| Tas | Cardiovascular | avg_weekly_apptemp | 0.459 (*) | -3.053 () | 0.076 () | -0.026 () |
| Tas | Cardiovascular | numb_days_over35 | -6.052 () | -200.036 (*) | 0.556 () | -2.63 () |
| Tas | Cardiovascular | numb_apptemp_hw_days | 0.704 (**) | 1.308 () | 0.157 (**) | -0.034 () |
| Tas | Cardiovascular | numb_temp_hw_days | 0.318 () | -5.304 () | -0.029 () | -0.04 () |
| Tas | Mental health | avg_weekly_max | -2.012 (**) | -4.549 (***) | -0.085 () | 0.104 () |
| Tas | Mental health | avg_weekly_apptemp | -1.605 (**) | -2.551 (**) | -0.19 (**) | -0.007 () |
| Tas | Mental health | numb_days_over35 | -41.133 (**) | -80.638 (***) | -1.939 () | 0.795 () |
| Tas | Mental health | numb_apptemp_hw_days | 0.19 () | 0.062 () | -0.015 () | 0.034 () |
| Tas | Mental health | numb_temp_hw_days | 0.733 () | -1.29 () | 0.004 () | 0.155 (*) |
| Tas | Respiratory | avg_weekly_max | -0.076 () | -3.674 (***) | -0.105 () | -0.564 (**) |
| Tas | Respiratory | avg_weekly_apptemp | 0.141 (**) | -0.665 () | 0.133 (*) | -0.152 () |
| Tas | Respiratory | numb_days_over35 | 1.929 () | -5.156 () | 1.288 () | 5.246 () |
| Tas | Respiratory | numb_apptemp_hw_days | 0.145 (**) | -0.098 () | 0.158 (**) | 0.108 () |
| Tas | Respiratory | numb_temp_hw_days | 0.02 () | -1.462 (*) | -0.066 () | -0.37 (*) |
| Tas | Total | avg_weekly_max | -2.788 (**) | -23.767 (***) | -0.097 () | -0.119 () |
| Tas | Total | avg_weekly_apptemp | -1.16 () | -6.269 () | -0.108 () | -0.47 (*) |
| Tas | Total | numb_days_over35 | -51.804 (**) | -285.829 (*) | -1.168 () | 8.489 () |
| Tas | Total | numb_apptemp_hw_days | 1.047 () | 1.273 () | 0.212 () | -0.088 () |
| Tas | Total | numb_temp_hw_days | 0.975 () | -8.056 () | 0.084 () | 0.007 () |
| Vic | Cardiovascular | avg_weekly_max | -0.176 () | -0.993 () | -0.053 (**) | -0.041 (*) |
| Vic | Cardiovascular | avg_weekly_apptemp | 0.421 (***) | -2.484 () | 0.033 () | 0.042 () |
| Vic | Cardiovascular | numb_days_over35 | -2.182 (***) | -12.409 () | -0.266 (**) | -0.256 (**) |
| Vic | Cardiovascular | numb_apptemp_hw_days | 0.86 (***) | -0.652 () | 0.082 (***) | 0.065 (**) |
| Vic | Cardiovascular | numb_temp_hw_days | 0.432 (***) | 0.603 () | 0.062 (**) | -0.016 () |
| Vic | Mental health | avg_weekly_max | 0.149 () | 0.061 () | 0.101 (***) | 0.103 (***) |
| Vic | Mental health | avg_weekly_apptemp | -0.926 (*) | -0.81 () | -0.002 () | 0.081 (***) |
| Vic | Mental health | numb_days_over35 | -6.571 (***) | -2.686 () | 0.126 () | 0.259 (***) |
| Vic | Mental health | numb_apptemp_hw_days | 1.617 (***) | 0.495 () | 0.03 () | 0.067 (***) |
| Vic | Mental health | numb_temp_hw_days | 2.313 (***) | 1.108 () | 0.068 (**) | 0.097 (***) |
| Vic | Respiratory | avg_weekly_max | -0.053 (**) | -1.788 (***) | -0.153 (***) | -0.63 (***) |
| Vic | Respiratory | avg_weekly_apptemp | 0.048 (**) | -0.568 (*) | 0.15 (***) | 0.063 () |
| Vic | Respiratory | numb_days_over35 | -0.324 (***) | -6.012 (***) | -0.552 (***) | -1.397 (***) |
| Vic | Respiratory | numb_apptemp_hw_days | 0.047 (*) | -1.006 (***) | 0.091 (***) | -0.072 () |
| Vic | Respiratory | numb_temp_hw_days | 0.029 () | -0.767 (**) | 0.04 () | -0.241 (***) |
| Vic | Total | avg_weekly_max | -0.108 () | -2.72 () | -0.116 (**) | -0.601 (***) |
| Vic | Total | avg_weekly_apptemp | -0.49 () | -3.863 () | 0.167 (***) | 0.112 () |
| Vic | Total | numb_days_over35 | -9.318 (***) | -21.107 (*) | -0.925 (***) | -2.063 (***) |
| Vic | Total | numb_apptemp_hw_days | 2.527 (***) | -1.164 () | 0.226 (***) | 0.019 () |
| Vic | Total | numb_temp_hw_days | 2.768 (***) | 0.943 () | 0.197 (***) | -0.133 () |
| WA | Cardiovascular | avg_weekly_max | -0.076 () | -16.601 (***) | -0.024 () | -0.086 () |
| WA | Cardiovascular | avg_weekly_apptemp | 0.204 (*) | -13.049 (***) | 0.044 () | 0.049 () |
| WA | Cardiovascular | numb_days_over35 | 0.433 (*) | -23.031 (***) | 0.136 (**) | -0.001 () |
| WA | Cardiovascular | numb_apptemp_hw_days | 0.19 () | -13.16 (***) | 0.005 () | -0.026 () |
| WA | Cardiovascular | numb_temp_hw_days | -0.005 () | -13.037 (***) | -0.015 () | -0.014 () |
| WA | Mental health | avg_weekly_max | -0.876 (*) | -3.3 (***) | 0.034 () | 0.026 () |
| WA | Mental health | avg_weekly_apptemp | 0.066 () | -2.047 (**) | 0.11 (***) | -0.021 () |
| WA | Mental health | numb_days_over35 | 1.031 () | -5.673 (***) | 0.165 (***) | -0.126 () |
| WA | Mental health | numb_apptemp_hw_days | -0.352 () | -2.74 (***) | 0.087 (***) | 0.035 () |
| WA | Mental health | numb_temp_hw_days | -0.237 () | -2.254 (**) | 0.014 () | 0.106 () |
| WA | Respiratory | avg_weekly_max | -0.14 (***) | -2.977 (***) | -0.065 () | -0.277 () |
| WA | Respiratory | avg_weekly_apptemp | -0.013 () | -0.734 (**) | -0.093 (**) | -0.655 (***) |
| WA | Respiratory | numb_days_over35 | 0.01 () | -0.671 () | -0.051 () | -1.008 (**) |
| WA | Respiratory | numb_apptemp_hw_days | -0.088 (**) | -2.252 (***) | -0.048 () | -0.46 (*) |
| WA | Respiratory | numb_temp_hw_days | -0.077 (*) | -1.161 (***) | 0.018 () | 0.258 () |
| WA | Total | avg_weekly_max | -0.991 (*) | -22.878 (***) | -0.137 (*) | -0.525 () |
| WA | Total | avg_weekly_apptemp | -0.041 () | -15.83 (***) | -0.053 () | -0.535 () |
| WA | Total | numb_days_over35 | 0.495 () | -29.376 (***) | 0.179 () | -0.844 () |
| WA | Total | numb_apptemp_hw_days | -0.649 () | -18.153 (***) | -0.095 () | -0.698 () |
| WA | Total | numb_temp_hw_days | -0.422 () | -16.452 (***) | -0.048 () | 0.274 () |
| ACT | Other | avg_weekly_max | |  | 0.018 () | 0.065 () |
| ACT | Other | avg_weekly_apptemp | |  | -0.05 () | -0.143 (*) |
| ACT | Other | numb_days_over35 | |  | -0.113 () | -0.649 (*) |
| ACT | Other | numb_apptemp_hw_days | |  | -0.054 () | -0.183 (*) |
| ACT | Other | numb_temp_hw_days | |  | 0.023 () | 0.188 (*) |
| NSW | Other | avg_weekly_max | |  | 0.014 () | 0.076 (***) |
| NSW | Other | avg_weekly_apptemp | |  | -0.027 () | -0.136 (***) |
| NSW | Other | numb_days_over35 | |  | 0.071 () | -0.258 (***) |
| NSW | Other | numb_apptemp_hw_days | |  | -0.016 () | -0.169 (***) |
| NSW | Other | numb_temp_hw_days | |  | -0.033 () | 0.073 (***) |
| NT | Other | avg_weekly_max | |  | 0.122 (**) | 0.705 (***) |
| NT | Other | avg_weekly_apptemp | |  | 0.058 () | 0.17 () |
| NT | Other | numb_days_over35 | |  | 0.194 (**) | 0.566 (***) |
| NT | Other | numb_apptemp_hw_days | |  | 0.04 () | 0.009 () |
| NT | Other | numb_temp_hw_days | |  | 0.038 () | -0.118 () |
| Qld | Other | avg_weekly_max | |  | -0.006 () | 0.208 (***) |
| Qld | Other | avg_weekly_apptemp | |  | -0.008 () | -0.141 (***) |
| Qld | Other | numb_days_over35 | |  | -0.161 (**) | 0.013 () |
| Qld | Other | numb_apptemp_hw_days | |  | -0.005 () | -0.155 (***) |
| Qld | Other | numb_temp_hw_days | |  | 0.021 () | 0.056 (**) |
| SA | Other | avg_weekly_max | |  | -0.005 () | -0.047 () |
| SA | Other | avg_weekly_apptemp | |  | -0.059 () | -0.094 (**) |
| SA | Other | numb_days_over35 | |  | -0.17 () | -0.413 (***) |
| SA | Other | numb_apptemp_hw_days | |  | -0.056 () | -0.107 (**) |
| SA | Other | numb_temp_hw_days | |  | 0.023 () | -0.035 () |
| Tas | Other | avg_weekly_max | |  | 0.127 () | 0.467 (***) |
| Tas | Other | avg_weekly_apptemp | |  | -0.085 () | -0.099 () |
| Tas | Other | numb_days_over35 | |  | -1.904 () | 3.258 () |
| Tas | Other | numb_apptemp_hw_days | |  | -0.044 () | -0.124 () |
| Tas | Other | numb_temp_hw_days | |  | 0.153 () | 0.229 (**) |
| Vic | Other | avg_weekly_max | |  | -0.01 () | -0.032 () |
| Vic | Other | avg_weekly_apptemp | |  | -0.014 () | -0.073 (***) |
| Vic | Other | numb_days_over35 | |  | -0.232 (***) | -0.669 (***) |
| Vic | Other | numb_apptemp_hw_days | |  | 0.023 () | -0.041 (*) |
| Vic | Other | numb_temp_hw_days | |  | 0.027 () | 0.028 () |
| WA | Other | avg_weekly_max | |  | -0.077 (*) | -0.077 () |
| WA | Other | avg_weekly_apptemp | |  | -0.108 (***) | -0.111 (*) |
| WA | Other | numb_days_over35 | |  | -0.078 () | -0.444 (***) |
| WA | Other | numb_apptemp_hw_days | |  | -0.122 (**) | -0.093 () |
| WA | Other | numb_temp_hw_days | |  | -0.067 () | 0.011 () |

### Supplementary Table 4: Climate zone-stratified model results

| climate zone group | condition | temp_var | MBS | PBS | Hospital | ED |
| --- | --- | --- | --- | --- | --- | --- |
| Hot Dry | Cardiovascular | avg_weekly_max | -0.262 () | -8.381 (***) | -0.174 (***) | 0.012 () |
| Hot Dry | Cardiovascular | avg_weekly_apptemp | 0.06 () | -1.029 () | -0.031 () | 0.029 () |
| Hot Dry | Cardiovascular | numb_days_over35 | -0.264 () | -27.198 (***) | 0.033 () | 0.047 () |
| Hot Dry | Cardiovascular | numb_apptemp_hw_days | 0.137 () | -0.447 () | 0.029 () | 0.068 () |
| Hot Dry | Cardiovascular | numb_temp_hw_days | 0.279 () | -8.08 (**) | -0.006 () | 0.004 () |
| Hot Dry | Mental health | avg_weekly_max | -1.082 (***) | -2.584 (***) | 0.094 (**) | 0.395 (***) |
| Hot Dry | Mental health | avg_weekly_apptemp | 0.605 (*) | 0.886 () | 0.115 (***) | 0.224 (**) |
| Hot Dry | Mental health | numb_days_over35 | -2.387 (***) | -9.833 (***) | 0.489 (***) | 0.771 (***) |
| Hot Dry | Mental health | numb_apptemp_hw_days | 0.556 () | 0.569 () | 0.112 (***) | 0.328 (***) |
| Hot Dry | Mental health | numb_temp_hw_days | 0.205 () | -1.619 (*) | 0.156 (***) | 0.178 () |
| Hot Dry | Respiratory | avg_weekly_max | -0.087 (*) | -2.98 (***) | -0.139 (**) | -0.492 (**) |
| Hot Dry | Respiratory | avg_weekly_apptemp | -0.032 () | -1.634 (***) | -0.14 (***) | -0.653 (***) |
| Hot Dry | Respiratory | numb_days_over35 | -0.309 (***) | -3.226 (***) | -0.208 (*) | -1.341 (***) |
| Hot Dry | Respiratory | numb_apptemp_hw_days | -0.013 () | -1.702 (***) | -0.046 () | -0.66 (**) |
| Hot Dry | Respiratory | numb_temp_hw_days | 0.023 () | -1.097 (**) | 0.14 (**) | -0.011 () |
| Hot Dry | Total | avg_weekly_max | -1.699 (***) | -13.945 (***) | -0.256 (**) | -0.261 () |
| Hot Dry | Total | avg_weekly_apptemp | 0.655 () | -1.776 () | -0.12 () | -0.418 () |
| Hot Dry | Total | numb_days_over35 | -3.229 (***) | -40.256 (***) | 0.263 () | -0.497 () |
| Hot Dry | Total | numb_apptemp_hw_days | 0.622 () | -1.58 () | 0.065 () | -0.288 () |
| Hot Dry | Total | numb_temp_hw_days | 0.428 () | -10.795 (**) | 0.284 (**) | 0.052 () |
| Hot Humid | Cardiovascular | avg_weekly_max | 0.245 () | -2.414 () | -0.055 (*) | 0.159 (**) |
| Hot Humid | Cardiovascular | avg_weekly_apptemp | 0.449 (***) | -3.355 (*) | 0.048 (**) | 0.081 (**) |
| Hot Humid | Cardiovascular | numb_days_over35 | 0.954 (***) | -2.578 () | 0.172 (***) | 0.192 (*) |
| Hot Humid | Cardiovascular | numb_apptemp_hw_days | 0.557 (***) | -5.221 (**) | 0.071 (***) | 0.068 (*) |
| Hot Humid | Cardiovascular | numb_temp_hw_days | 0.479 (***) | -0.945 () | 0.052 (**) | 0.011 () |
| Hot Humid | Mental health | avg_weekly_max | 0.855 () | -0.368 () | 0.042 () | 0.189 (***) |
| Hot Humid | Mental health | avg_weekly_apptemp | -0.04 () | -0.452 () | 0.093 (***) | 0.081 (***) |
| Hot Humid | Mental health | numb_days_over35 | 0.205 () | -5.182 (***) | 0.241 (***) | 0.064 () |
| Hot Humid | Mental health | numb_apptemp_hw_days | 0.958 (**) | -0.813 () | 0.105 (***) | 0.073 (**) |
| Hot Humid | Mental health | numb_temp_hw_days | 0.782 (*) | 0.287 () | 0.077 (**) | 0.049 () |
| Hot Humid | Respiratory | avg_weekly_max | -0.039 () | -4.436 (***) | -0.485 (***) | -0.774 (***) |
| Hot Humid | Respiratory | avg_weekly_apptemp | 0.043 (*) | -1.417 (***) | 0.004 () | -0.017 () |
| Hot Humid | Respiratory | numb_days_over35 | 0.338 (***) | 1.504 (**) | -0.617 (***) | -1.074 (***) |
| Hot Humid | Respiratory | numb_apptemp_hw_days | 0.065 (**) | -1.818 (***) | -0.004 () | -0.288 (**) |
| Hot Humid | Respiratory | numb_temp_hw_days | 0.095 (***) | -0.868 (***) | 0.059 (*) | 0.116 () |
| Hot Humid | Total | avg_weekly_max | 0.411 () | -7.219 (*) | -0.529 (***) | -0.38 () |
| Hot Humid | Total | avg_weekly_apptemp | 0.407 () | -5.223 (**) | 0.143 (**) | -0.021 () |
| Hot Humid | Total | numb_days_over35 | 1.159 () | -6.256 () | -0.297 (*) | -0.301 () |
| Hot Humid | Total | numb_apptemp_hw_days | 1.414 (**) | -7.852 (***) | 0.165 (**) | -0.391 (**) |
| Hot Humid | Total | numb_temp_hw_days | 1.011 (*) | -1.527 () | 0.185 (***) | 0.023 () |
| Temperate/Cold | Cardiovascular | avg_weekly_max | -0.085 () | -0.813 () | -0.021 () | 0.011 () |
| Temperate/Cold | Cardiovascular | avg_weekly_apptemp | 0.039 () | -4.328 (***) | 0.002 () | -0.013 () |
| Temperate/Cold | Cardiovascular | numb_days_over35 | -0.638 (**) | -6.295 () | -0.026 () | 0.001 () |
| Temperate/Cold | Cardiovascular | numb_apptemp_hw_days | 0.441 (***) | -4.022 (***) | 0.032 (**) | 0.015 () |
| Temperate/Cold | Cardiovascular | numb_temp_hw_days | 0.063 () | -3.261 (**) | 0.002 () | -0.011 () |
| Temperate/Cold | Mental health | avg_weekly_max | -0.924 (***) | -0.63 () | 0.113 (***) | 0.096 (***) |
| Temperate/Cold | Mental health | avg_weekly_apptemp | 0.327 (*) | -0.065 () | -0.017 () | 0.091 (***) |
| Temperate/Cold | Mental health | numb_days_over35 | -3.311 (***) | -4.257 (**) | 0.305 (***) | 0.017 () |
| Temperate/Cold | Mental health | numb_apptemp_hw_days | 1.448 (***) | 0.162 () | 0.056 (***) | 0.095 (***) |
| Temperate/Cold | Mental health | numb_temp_hw_days | 0.376 () | 0.222 () | 0.043 (**) | 0.117 (***) |
| Temperate/Cold | Respiratory | avg_weekly_max | -0.011 () | -1.089 (***) | -0.164 (***) | -0.327 (***) |
| Temperate/Cold | Respiratory | avg_weekly_apptemp | -0.029 (*) | -1.813 (***) | 0.08 (***) | -0.185 (***) |
| Temperate/Cold | Respiratory | numb_days_over35 | -0.15 (**) | -4.005 (***) | -0.269 (***) | -0.082 () |
| Temperate/Cold | Respiratory | numb_apptemp_hw_days | 0.023 () | -1.898 (***) | 0.011 () | -0.26 (***) |
| Temperate/Cold | Respiratory | numb_temp_hw_days | -0.002 () | -0.998 (***) | 0.006 () | -0.032 () |
| Temperate/Cold | Total | avg_weekly_max | -0.948 (***) | -2.532 () | -0.058 (*) | -0.161 (**) |
| Temperate/Cold | Total | avg_weekly_apptemp | 0.318 () | -6.205 (***) | 0.055 (*) | -0.221 (***) |
| Temperate/Cold | Total | numb_days_over35 | -3.772 (***) | -14.557 (**) | -0.046 () | -0.533 (**) |
| Temperate/Cold | Total | numb_apptemp_hw_days | 1.916 (***) | -5.759 (***) | 0.082 (**) | -0.262 (***) |
| Temperate/Cold | Total | numb_temp_hw_days | 0.418 () | -4.037 (*) | 0.063 (*) | 0.146 (*) |
| Hot Dry | Other | avg_weekly_max |  |  | -0.036 () | -0.109 (**) |
| Hot Dry | Other | avg_weekly_apptemp |  |  | -0.063 (*) | 0.014 () |
| Hot Dry | Other | numb_days_over35 |  |  | -0.057 () | 0.026 () |
| Hot Dry | Other | numb_apptemp_hw_days |  |  | -0.027 () | -0.04 () |
| Hot Dry | Other | numb_temp_hw_days |  |  | -0.006 () | 0.008 () |
| Hot Humid | Other | avg_weekly_max |  |  | -0.031 () | 0.119 (**) |
| Hot Humid | Other | avg_weekly_apptemp |  |  | -0.002 () | -0.152 (***) |
| Hot Humid | Other | numb_days_over35 |  |  | -0.1 () | -0.096 () |
| Hot Humid | Other | numb_apptemp_hw_days |  |  | -0.006 () | -0.162 (***) |
| Hot Humid | Other | numb_temp_hw_days |  |  | -0.002 () | -0.015 () |
| Temperate/Cold | Other | avg_weekly_max |  |  | 0.005 () | 0.056 (***) |
| Temperate/Cold | Other | avg_weekly_apptemp |  |  | -0.022 (*) | -0.112 (***) |
| Temperate/Cold | Other | numb_days_over35 |  |  | -0.106 (*) | -0.487 (***) |
| Temperate/Cold | Other | numb_apptemp_hw_days |  |  | -0.016 () | -0.121 (***) |
| Temperate/Cold | Other | numb_temp_hw_days |  |  | 0.008 () | 0.078 (***) |

### Supplementary Table 5: Interaction term results: IRSAD relative rank

| **condition** | **temp_var** | **MBS** | **PBS** | **Hospital** | **ED** |
| --- | --- | --- | --- | --- | --- |
| Cardiovascular | numb_apptemp_hw_days | -0.602 (***) | -35.69 (***) | -0.143 (***) | -0.063 () |
| Cardiovascular | numb_temp_hw_days | -0.831 (***) | -15.76 (***) | -0.103 (**) | -0.058 () |
| Mental health | numb_apptemp_hw_days | 8.533 (***) | -9.345 (***) | 0.306 (***) | -0.062 () |
| Mental health | numb_temp_hw_days | 3.582 (***) | -4.341 (***) | 0.212 (***) | -0.163 (**) |
| Other | numb_apptemp_hw_days |  |  | -0.024 () | -0.018 () |
| Other | numb_temp_hw_days |  |  | -0.072 (*) | 0.045 () |
| Respiratory | numb_apptemp_hw_days | -0.195 (***) | -3.624 (***) | -0.07 (.) | 0.242 (.) |
| Respiratory | numb_temp_hw_days | -0.073 (.) | -3.423 (***) | -0.379 (***) | -0.393 (**) |
| Total | numb_apptemp_hw_days | 8.676 (***) | -48.659 (***) | 0.048 () | 0.165 () |
| Total | numb_temp_hw_days | 3.565 (***) | -23.524 (***) | -0.369 (***) | -0.501 (**) |

### Supplementary Table 6: Interaction term results: Outdoor work index

| **condition** | **temp_var** | **MBS** | **PBS** | **Hospital** | **ED** |
| --- | --- | --- | --- | --- | --- |
| Cardiovascular | numb_apptemp_hw_days | 0.012 () | 1.932 (***) | 0.009 (***) | 0.002 () |
| Cardiovascular | numb_temp_hw_days | 0.019 (*) | 0.949 (***) | 0.006 (***) | 0.002 () |
| Mental health | numb_apptemp_hw_days | -0.52 (***) | 0.471 (***) | -0.018 (***) | 0.001 () |
| Mental health | numb_temp_hw_days | -0.247 (***) | 0.218 (***) | -0.017 (***) | 0.008 (**) |
| Other | numb_apptemp_hw_days |  |  | -0.004 (*) | 0.003 () |
| Other | numb_temp_hw_days |  |  | -0.002 () | -0.005 (**) |
| Respiratory | numb_apptemp_hw_days | 0.008 (***) | 0.2 (***) | 0.004 (.) | -0.015 (*) |
| Respiratory | numb_temp_hw_days | 0.002 () | 0.169 (***) | 0.019 (***) | 0.025 (***) |
| Total | numb_apptemp_hw_days | -0.573 (***) | 2.603 (***) | -0.009 (*) | -0.018 (.) |
| Total | numb_temp_hw_days | -0.301 (***) | 1.336 (***) | 0.008 (.) | 0.021 (*) |

### Supplementary Table 7: Chronic exposure model results

| clincal group | term_label | MBS | PBS | Hospital | ED |
| --- | --- | --- | --- | --- | --- |
| Cardiovascular | % Aged 65+ | 0.91 (***) | 0.71 (***) | 0.71 (***) | -0.1 () |
| Cardiovascular | % Chronic Illness | -0.37 () | 0.12 () | 0.12 () | 0.8 (***) |
| Cardiovascular | IRSAD Rank | -0.34 (*) | -0.27 (***) | -0.09 () | -0.2 () |
| Cardiovascular | Long-term Max Temp | 0.09 () | -0.12 (**) | 0.22 (**) | 0.4 (***) |
| Cardiovascular | Outdoor Work Index | -0.57 (**) | -0.08 () | 0.06 () | -0.38 () |
| Cardiovascular | Remoteness (MMM7) | -0.13 () | -0.17 (*) | 0.1 () | 0.22 () |
| Cardiovascular | accessibility_proxy | -0.26 (*) | -0.06 () | 0.04 () | -0.07 () |
| Mental health | % Aged 65+ | 0.09 () | 0.11 () | -0.19 () | -0.36 (*) |
| Mental health | % Chronic Illness | 0.28 () | 0.9 (***) | 0.54 (*) | 0.41 () |
| Mental health | IRSAD Rank | 0.29 (*) | -0.01 () | 0.43 (*) | -0.19 () |
| Mental health | Long-term Max Temp | -0.1 () | -0.01 () | 0.17 () | 0.25 (*) |
| Mental health | Outdoor Work Index | -0.52 (**) | 0.15 () | -0.74 (**) | -0.58 (**) |
| Mental health | Remoteness (MMM7) | -0.13 () | -0.26 (***) | 0.41 (*) | 0.73 (***) |
| Mental health | accessibility_proxy | 0.09 () | 0.21 (***) | -0.03 () | 0.12 () |
| Other | % Aged 65+ |  |  | 0.91 (***) | -0.21 () |
| Other | % Chronic Illness |  |  | -0.22 () | 0.58 (*) |
| Other | IRSAD Rank |  |  | -0.2 () | 0.07 () |
| Other | Long-term Max Temp |  |  | 0.15 () | 0.41 (***) |
| Other | Outdoor Work Index |  |  | -0.35 () | -0.45 (*) |
| Other | Remoteness (MMM7) |  |  | 0.23 () | 0.63 (***) |
| Other | accessibility_proxy |  |  | 0.06 () | -0.06 () |
| Respiratory | % Aged 65+ | 0.54 (**) | 0.29 (**) | 0 () | -0.28 () |
| Respiratory | % Chronic Illness | 0.28 () | 0.42 (***) | 0.05 () | 0.39 (*) |
| Respiratory | IRSAD Rank | -0.03 () | -0.42 (***) | -0.41 (**) | -0.19 () |
| Respiratory | Long-term Max Temp | 0.56 (***) | -0.15 (*) | 0.27 (***) | 0.29 (**) |
| Respiratory | Outdoor Work Index | -0.19 () | -0.21 () | -0.15 () | -0.58 (**) |
| Respiratory | Remoteness (MMM7) | -0.3 () | -0.14 () | 0.52 (***) | 0.84 (***) |
| Respiratory | accessibility_proxy | -0.06 () | -0.08 () | -0.01 () | -0.11 () |
| Total | % Aged 65+ | 0.29 (*) | 0.59 (***) | 0.55 (***) | -0.28 () |
| Total | % Chronic Illness | 0.19 () | 0.3 (***) | 0.29 () | 0.59 (**) |
| Total | IRSAD Rank | 0.19 () | -0.24 (***) | -0.06 () | -0.19 () |
| Total | Long-term Max Temp | -0.05 () | -0.11 (**) | 0.39 (***) | 0.37 (***) |
| Total | Outdoor Work Index | -0.6 (***) | -0.05 () | -0.54 (**) | -0.63 (**) |
| Total | Remoteness (MMM7) | -0.17 () | -0.19 (**) | 0.55 (***) | 0.76 (***) |
| Total | accessibility_proxy | 0.03 () | -0.01 () | 0.03 () | -0.05 () |

### Supplementary Table 8: Chronic exposure model results, 5 versus 20 years

| service | clincal group | temp_type | 20 years | Recent 5 years |
| --- | --- | --- | --- | --- |
| ED | Cardiovascular | Apparent | 0.986 (***) | 0.842 (**) |
| ED | Cardiovascular | Dry | 1.058 (***) | 0.993 (**) |
| ED | Mental health | Apparent | 0.134 () | 0.263 () |
| ED | Mental health | Dry | 0.635 () | 0.77 (*) |
| ED | Other | Apparent | 0.729 (***) | 0.771 (***) |
| ED | Other | Dry | 0.689 (**) | 0.808 (***) |
| ED | Respiratory | Apparent | 0.989 () | 1.387 (*) |
| ED | Respiratory | Dry | 1.778 (*) | 2.226 (**) |
| ED | Total | Apparent | 2.824 (*) | 3.311 (*) |
| ED | Total | Dry | 4.193 (**) | 4.93 (***) |
| Hospital | Cardiovascular | Apparent | 0.397 (*) | 0.908 (***) |
| Hospital | Cardiovascular | Dry | 0.662 (**) | 1.094 (***) |
| Hospital | Mental health | Apparent | 0.617 () | 1.741 (***) |
| Hospital | Mental health | Dry | 0.324 () | 1.203 (*) |
| Hospital | Other | Apparent | 0.393 (*) | 0.502 () |
| Hospital | Other | Dry | 0.238 () | 0.359 () |
| Hospital | Respiratory | Apparent | 0.208 () | 0.943 (***) |
| Hospital | Respiratory | Dry | 0.673 (**) | 1.247 (***) |
| Hospital | Total | Apparent | 1.662 (**) | 4.105 (***) |
| Hospital | Total | Dry | 2.033 (***) | 3.94 (***) |
| MBS | Cardiovascular | Apparent | -0.043 () | 2.695 (*) |
| MBS | Cardiovascular | Dry | 0.495 () | 4.277 (**) |
| MBS | Mental health | Apparent | -2.065 () | -0.227 () |
| MBS | Mental health | Dry | -9.132 () | -8.903 () |
| MBS | Respiratory | Apparent | 1.626 (***) | 1.395 (***) |
| MBS | Respiratory | Dry | 1.68 (***) | 1.48 (***) |
| MBS | Total | Apparent | -0.633 () | 3.879 () |
| MBS | Total | Dry | -7.092 () | -3.064 () |
| PBS | Cardiovascular | Apparent | -75.335 (**) | -69.65 (**) |
| PBS | Cardiovascular | Dry | -69.502 (**) | -55.883 (*) |
| PBS | Mental health | Apparent | 1.993 () | 9.119 () |
| PBS | Mental health | Dry | -0.337 () | 4.216 () |
| PBS | Respiratory | Apparent | -8.662 (**) | -8.453 (**) |
| PBS | Respiratory | Dry | -8.112 (**) | -7.844 (**) |
| PBS | Total | Apparent | -82.003 (**) | -68.984 (**) |
| PBS | Total | Dry | -77.951 (**) | -59.511 (*) |

Supplementary Table 6: Chronic exposure model results, climactic zones

| service | clincal group | zone | estimate_ci | sig | p.value |
| --- | --- | --- | --- | --- | --- |
| ED | Cardiovascular | Hot Dry | 0.85 (-3.49, 5.19) |  | 0.737630583 |
| ED | Cardiovascular | Hot Humid | 0.34 (-0.28, 0.95) |  | 0.300197223 |
| ED | Cardiovascular | Temperate/Cold | 0.00 (-0.20, 0.21) |  | 0.966372043 |
| ED | Mental health | Hot Dry | 1.05 (-0.48, 2.58) |  | 0.31171075 |
| ED | Mental health | Hot Humid | 0.24 (-0.50, 0.98) |  | 0.533512563 |
| ED | Mental health | Temperate/Cold | 0.35 (0.11, 0.60) | ** | 0.00716882 |
| ED | Other | Hot Dry | 0.53 (-1.95, 3.00) |  | 0.715923259 |
| ED | Other | Hot Humid | 0.12 (-0.76, 0.99) |  | 0.800481932 |
| ED | Other | Temperate/Cold | 0.14 (-0.12, 0.40) |  | 0.30112698 |
| ED | Respiratory | Hot Dry | 0.53 (-2.99, 4.05) |  | 0.796224732 |
| ED | Respiratory | Hot Humid | 0.02 (-0.68, 0.72) |  | 0.958604716 |
| ED | Respiratory | Temperate/Cold | 0.26 (0.03, 0.50) | * | 0.032980264 |
| ED | Total | Hot Dry | 0.70 (-2.09, 3.50) |  | 0.671575405 |
| ED | Total | Hot Humid | 0.17 (-0.59, 0.94) |  | 0.666252225 |
| ED | Total | Temperate/Cold | 0.25 (0.02, 0.48) | * | 0.036114096 |
| Hospital | Cardiovascular | Hot Dry | 0.26 (-3.97, 4.49) |  | 0.914755895 |
| Hospital | Cardiovascular | Hot Humid | 0.19 (-0.11, 0.49) |  | 0.23992644 |
| Hospital | Cardiovascular | Temperate/Cold | 0.04 (-0.10, 0.17) |  | 0.579750126 |
| Hospital | Mental health | Hot Dry | 2.04 (0.83, 3.24) |  | 0.080318016 |
| Hospital | Mental health | Hot Humid | -0.62 (-1.19, -0.05) |  | 0.053452351 |
| Hospital | Mental health | Temperate/Cold | 0.15 (-0.10, 0.40) |  | 0.243121127 |
| Hospital | Other | Hot Dry | -0.16 (-3.00, 2.68) |  | 0.923301487 |
| Hospital | Other | Hot Humid | -0.48 (-0.93, -0.02) |  | 0.060360468 |
| Hospital | Other | Temperate/Cold | -0.02 (-0.24, 0.21) |  | 0.871364539 |
| Hospital | Respiratory | Hot Dry | 0.40 (-1.70, 2.50) |  | 0.746492568 |
| Hospital | Respiratory | Hot Humid | -0.10 (-0.62, 0.42) |  | 0.708432068 |
| Hospital | Respiratory | Temperate/Cold | 0.12 (-0.07, 0.32) |  | 0.219976708 |
| Hospital | Total | Hot Dry | 0.86 (-2.97, 4.70) |  | 0.702550071 |
| Hospital | Total | Hot Humid | -0.56 (-1.19, 0.07) |  | 0.106286615 |
| Hospital | Total | Temperate/Cold | 0.16 (-0.01, 0.32) |  | 0.072423601 |
| MBS | Cardiovascular | Hot Dry | -0.50 (-2.58, 1.57) |  | 0.681281769 |
| MBS | Cardiovascular | Hot Humid | -0.37 (-0.68, -0.06) | * | 0.034618245 |
| MBS | Cardiovascular | Temperate/Cold | 0.17 (-0.09, 0.42) |  | 0.212765851 |
| MBS | Mental health | Hot Dry | -1.00 (-3.88, 1.88) |  | 0.565945722 |
| MBS | Mental health | Hot Humid | -0.78 (-1.07, -0.50) | *** | 0.00012935 |
| MBS | Mental health | Temperate/Cold | -0.26 (-0.44, -0.09) | ** | 0.004771838 |
| MBS | Respiratory | Hot Dry | 0.31 (-1.92, 2.54) |  | 0.811796266 |
| MBS | Respiratory | Hot Humid | -0.46 (-0.88, -0.05) | * | 0.04769209 |
| MBS | Respiratory | Temperate/Cold | 0.53 (0.28, 0.78) | *** | 0.000141369 |
| MBS | Total | Hot Dry | -0.87 (-3.17, 1.44) |  | 0.538100376 |
| MBS | Total | Hot Humid | -0.75 (-1.00, -0.50) | *** | 5.80108E-05 |
| MBS | Total | Temperate/Cold | -0.18 (-0.38, 0.02) |  | 0.080533309 |
| PBS | Cardiovascular | Hot Dry | -0.01 (-0.40, 0.37) |  | 0.94760615 |
| PBS | Cardiovascular | Hot Humid | -0.18 (-0.43, 0.08) |  | 0.193878635 |
| PBS | Cardiovascular | Temperate/Cold | 0.02 (-0.07, 0.11) |  | 0.657134484 |
| PBS | Mental health | Hot Dry | 0.07 (-0.21, 0.36) |  | 0.666891127 |
| PBS | Mental health | Hot Humid | -0.24 (-0.45, -0.04) | * | 0.036836833 |
| PBS | Mental health | Temperate/Cold | -0.02 (-0.10, 0.06) |  | 0.696221609 |
| PBS | Respiratory | Hot Dry | -0.01 (-1.18, 1.16) |  | 0.98976332 |
| PBS | Respiratory | Hot Humid | -0.24 (-0.50, 0.03) |  | 0.100060089 |
| PBS | Respiratory | Temperate/Cold | -0.04 (-0.17, 0.09) |  | 0.560918048 |
| PBS | Total | Hot Dry | 0.00 (-0.24, 0.25) |  | 0.994293606 |
| PBS | Total | Hot Humid | -0.19 (-0.41, 0.02) |  | 0.105481383 |
| PBS | Total | Temperate/Cold | 0.01 (-0.07, 0.09) |  | 0.806971628 |
